# Multi-Contrast Deep-Silicon Photon-Counting CT for Post-Embolization Imaging: A Quantitative Phantom Study

**DOI:** 10.64898/2026.10.01.26364507

**Authors:** Navid Azimi, Thomas Wesley Holmes, Arnaud R. Brian-Choux, Zhye Yin, Dominic Crotty, Nariman Nezami, Amir Pourmorteza

## Abstract

**Purpose:** To evaluate whether eight-bin deep-silicon photon-counting CT can quantify iodine and gadolinium separately and distinguish radiopaque embolic beads from vascular contrast in tumor phantoms.

**Materials and Methods:** In this phantom study, two-input decomposition of scanner-generated iodine and water images was compared with three- and five-material decomposition of five reconstructed energy-bin images in 56 common test vials, including nine simulated-blood preparations. One tantalum and one bismuth vial were also tested with five-material decomposition. A separately calibrated two-input method using scanner-generated 70-keV and iodine images was applied to four static 3D-printed tumor vascular models: iodine-only and gadolinium-only controls and two models with iodine-containing beads and gadolinium-filled channels. Concentration errors were summarized by bias and root mean square error (RMSE).

**Results:** For iodine, RMSE was 0.40, 0.73, and 1.12 mg I/mL with two-input, three-material, and five-material decomposition, respectively; corresponding gadolinium RMSEs were 0.57, 0.55, and 0.46 mg Gd/mL. Two-input biases were +0.01 mg I/mL and +0.29 mg Gd/mL. Five-material decomposition distinguished tantalum and bismuth in the separate test vials. Beads and vascular gadolinium were spatially separated in the tumor models. In the two bead-containing models, gadolinium measured 32.56 ± 8.02 and 37.02 ± 7.93 mg Gd/mL (mean ± spatial SD), compared with the prepared 35.00 mg Gd/mL.

**Conclusion:** Deep-silicon photon-counting CT separately visualized radiopaque embolic beads and quantified vascular gadolinium from a single acquisition. Recalibrated scanner-generated images supported quantitative dual-contrast imaging, while research energy-bin decomposition distinguished additional materials.

## Introduction

Photon-counting CT (PCCT) uses energy-resolving detectors to provide spectral information and improved spatial resolution compared with conventional energy-integrating detector CT (Willemink et al., 2018). Current commercial cadmium-telluride (CdTe)-based PCCT uses separate spectral and ultrahigh-resolution acquisition modes, requiring the mode to be selected before scanning (Mergen et al., 2025). In contrast, the new deep-silicon (dSi) PCCT system acquires eight energy bins at ultrahigh resolution in every scan, making both capabilities available from the same acquisition.

A previous dSi-PCCT study separated iodine and gadolinium while preserving ultrahigh-resolution characterization of calcified coronary plaques and stenoses (Holmes et al., 2024). An earlier four-bin CdTe prototype separated three contrast materials, iodine, gadolinium, and bismuth, in vivo (Symons et al., 2017). Here, we demonstrate the potential value of dSi-PCCT for quantitative decomposition of up to five materials. The five-material method uses research energy-bin images, whereas a complementary two-input method recalibrates the scanner’s FDA-cleared iodine and water maps for iodine/gadolinium separation.

Multimaterial imaging may be particularly useful in cancer treatment, where the distribution of radiopaque embolic material and persistent tumor enhancement provide different information about the treated region. Radiopaque beads can show where treatment was deposited (Reicher et al., 2019), whereas residual enhancement may indicate persistent viable tumor (Aslam et al., 2024). When the embolic material and intravascular contrast contain the same element, their signals cannot be separated on conventional CT. Subtraction imaging requires alignment across scans, which can be disrupted by motion or interval changes (Okumura et al., 2007); cardiac-induced abdominal-organ motion is one such source (Holmes et al., 2024). Distinguishing the two materials within a single acquisition could show embolic material deposition and residual enhancement independently. The purpose of this study was to evaluate quantitative multimaterial imaging with dSi-PCCT in phantoms containing multiple contrast agents and 3D-printed tumor models.

## Materials and Methods

### Study design

This phantom study evaluated two complementary approaches to multimaterial imaging with an eight-bin dSi-PCCT system. Energy-bin images were used to separate and quantify up to five materials, whereas scanner-generated iodine and water images were used for iodine/gadolinium separation. The latter approach was also applied to 3D-printed tumor models to distinguish radiopaque embolic material from vascular contrast.

This study was supported by a sponsored research agreement between [redacted] University and GE HealthCare. Authors who were neither employees of nor consultants for GE HealthCare had complete control of the data and analyses and the decision to submit the manuscript for publication.

### Test Objects

#### Multi-vial phantom

A 200-mm-diameter modular phantom contained 137 removable 2-mL microcentrifuge vials within a 3D-printed housing filled with 20% synthetic ballistic gel (Clear Ballistics, Greenville, SC) (Figure 1). The phantom contained iodine, gadolinium, and iodine–gadolinium preparations diluted in either water or simulated blood, together with water and simulated-blood references. Iodine and gadolinium solutions were prepared from iohexol (Omnipaque 350; GE HealthCare, Chicago, IL) and gadobutrol (Gadavist; Bayer HealthCare Pharmaceuticals, Whippany, NJ), respectively. Simulated blood was prepared by diluting ferrous sulfate oral drops (Fer-In-Sol Iron Drops; Lannett Company, Trevose, PA) in water to 3.75 mg Fe/mL, selected to produce approximately 55 HU at 120 kVp on an energy-integrating detector CT scanner.

**Figure 1.**
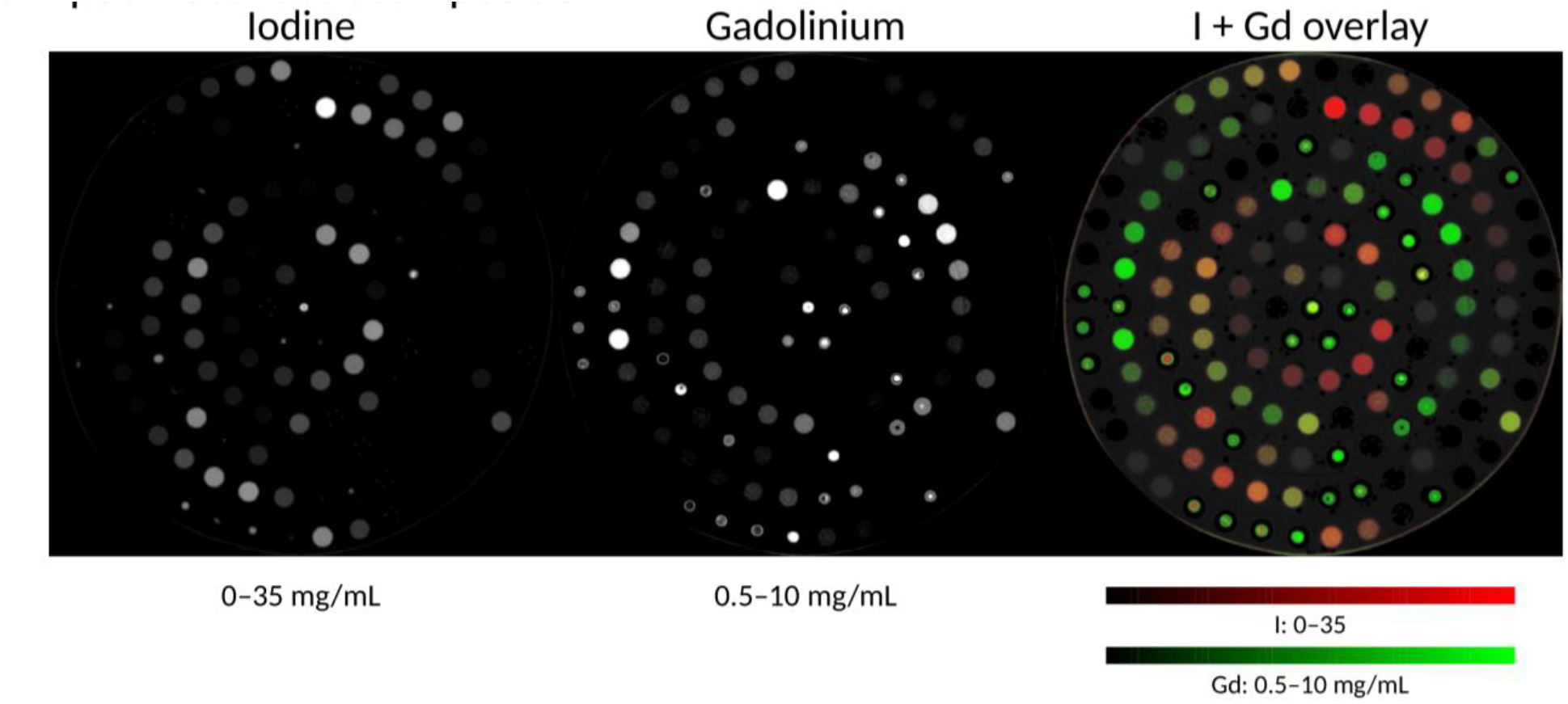
Iodine and gadolinium separation in the multi-vial phantom. From left to right: iodine map, gadolinium map, and combined iodine (red) and gadolinium (green) overlay on the 70-keV image. Iodine and gadolinium maps were generated by two-input decomposition of the scanner-generated iodine and water images. Images represent the mean of five contiguous axial slices. Display ranges are 0–35 mg/mL for iodine and 0.5–10 mg/mL for gadolinium.The 70-keV background is displayed with a window width/level of 1800/600 HU. Gd = gadolinium; HU = Hounsfield units.

Two vials contained TaCZ, a carboxybetaine zwitterionic-coated tantalum oxide nanoparticle preparation, diluted in water to 10 mg Ta/mL (FitzGerald et al., 2016). Two additional vials contained undiluted regular-strength bismuth subsalicylate suspension (Pepto-Bismol; The Procter & Gamble Manufacturing Company, Cincinnati, OH), corresponding to 10.1 mg Bi/mL.

#### Tumor vascular models

Four tumor vascular models incorporating branched vascular channels and focal luminal narrowing were printed by stereolithography (Mono 4K; Anycubic, Shenzhen, China) using clear green resin (ABS-Like Resin Pro 2; Anycubic) (Figure 2). Two models were prepared with 70–150 µm radiopaque embolic beads (LC Bead LUMI; Boston Scientific Corporation, Marlborough, MA). A 10-mL prefilled bead vial was shaken, and approximately 2 mL of its contents was injected into each model using a 5-mL syringe. The bead suspension was used without added saline, soluble iodine, or drug. Each bead-containing model was then flushed with saline, filled with 35 mg Gd/mL gadobutrol, and sealed. The two control models were filled with either 50 mg I/mL iohexol or 35 mg Gd/mL gadobutrol without beads.

**Figure 2.**
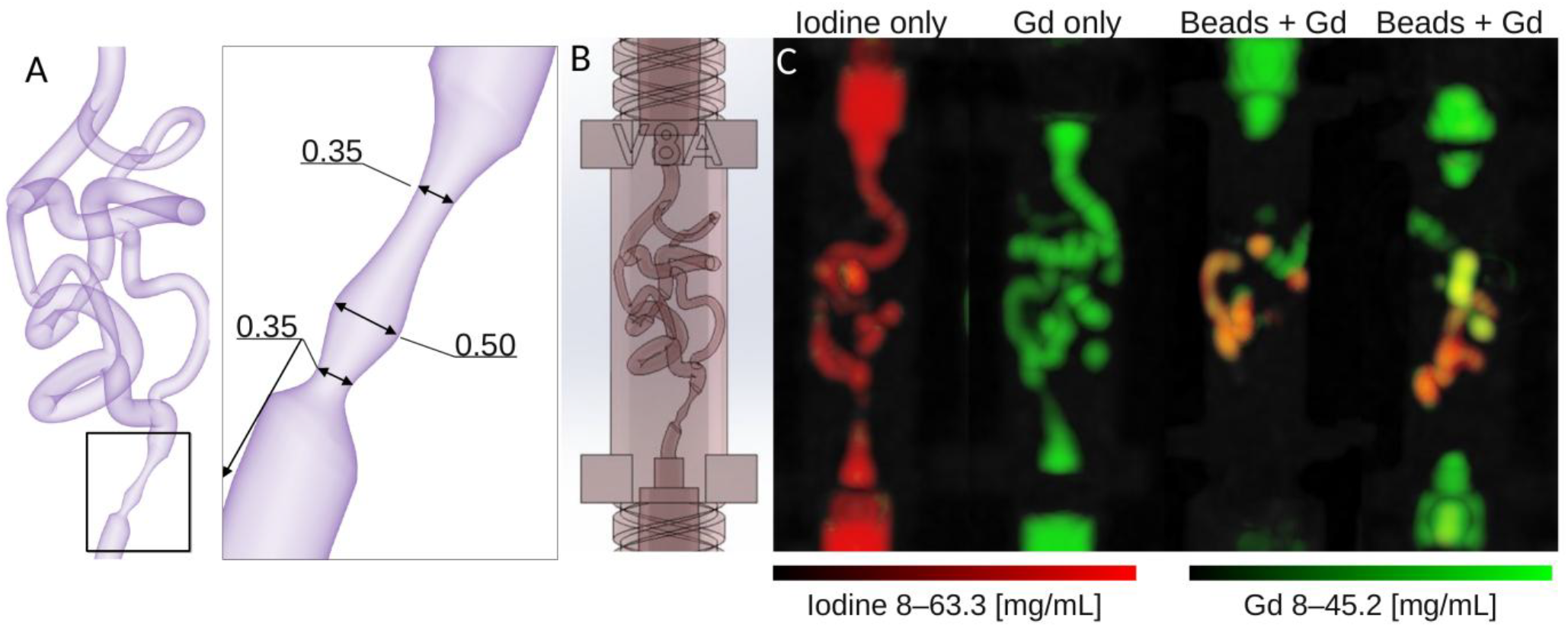
Separation of radiopaque embolic beads from vascular gadolinium in 3D-printed tumor models. (A) Three-dimensional rendering of the vascular network and an enlarged view of the focal narrowings, with lumen diameters in millimeters. (B) Computer-aided design of the vascular network within the insert housing. (C) Maximum-intensity projections showing iodine in red and gadolinium in green over the 70-keV image. From left to right: iodine-only control, gadolinium-only control, and two bead-containing models filled with gadolinium. Material maps were generated by two-input decomposition of the scanner-generated 70-keV and iodine images. Display ranges are 8–63.3 mg/mL for iodine and 8–45.2 mg/mL for gadolinium. Gd = gadolinium.

For scanning, the models were placed in a water-filled bag within the central opening of a custom-made simulated adipose tissue-equivalent plastic ring. The ring had a 20-cm outer diameter, a 9-cm central opening, and mean attenuation of approximately ™80 HU at 120 kVp.

### CT System

All scans were performed at the GE HealthCare facility in Waukesha, Wisconsin, using a deep-silicon photon-counting CT system (Photonova Spectra; GE HealthCare, Waukesha, WI). The system acquires eight energy bins at ultra-high-resolution (UHR) during every scan.

### Acquisition and Reconstruction Parameters

All acquisitions used 120 kVp and a 1-second rotation time. The multi-vial phantom was scanned at 300 mAs, and the tumor phantom was scanned at 255 mAs. Three lower energy thresholds were used to capture Compton effects, and five adjustable thresholds were set to 33, 44, 52, 60, and 80 keV.

All images were reconstructed using filtered backprojection with a 1024 × 1024 matrix. Virtual monoenergetic images (VMI) and iodine/water maps were generated on the scanner. Energy-bin images were generated using an offline research tool and did not include all detailed calibration and artifact-correction procedures applied to the scanner-generated images. Multi-vial phantom images were reconstructed with a 200-mm FOV and 0.195 × 0.195 × 0.417-mm^3^ voxels. Tumor phantom images were reconstructed with a 100-mm FOV and 0.098 × 0.098 × 0.417-mm^3^ voxels.

### Quantitative Image Analysis

For the multi-vial phantom, images were averaged over >2 mm in the z direction. Circular regions of interest (ROIs) >2.5 mm in diameter were manually placed within each vial using MATLAB (MathWorks, Natick, MA). Mean ROI measurements were used for calibration and testing.

Material decomposition followed previously described linear calibration methods (Symons et al., 2017; Holmes et al., 2024). Three-material decomposition used five energy-bin images to estimate iodine, gadolinium, and simulated blood and was calibrated with 22 vials. Five-material decomposition used the same image inputs to estimate iodine, gadolinium, simulated blood, tantalum, and bismuth and was calibrated with 24 vials. Two-input decomposition used scanner-generated iodine and water images to estimate iodine and gadolinium and was calibrated with 10 vials. Water-reference vials were used for baseline correction in each method.

All three methods were evaluated in the same 56 test vials: 47 containing iodine, gadolinium, or both in water and nine containing simulated blood alone or mixed with iodine or gadolinium. One additional tantalum vial and one additional bismuth vial were evaluated with five-material decomposition.

For the tumor vascular models, two-input decomposition was applied voxel-wise to the scanner-generated images. Maximum-intensity projections and combined iodine/gadolinium overlays were generated for display. Iodine and gadolinium concentrations were measured using manually selected three-dimensional ROIs in fluid-filled regions upstream or downstream from the tumor.

## Statistical Analysis

### Statistical analyses were performed in MATLAB

Each test vial contributed one mean concentration measurement. Concentration error was calculated as estimated minus reference concentration. Bias was the mean error, and root-mean-square error (RMSE) was calculated from the vial-level errors. Approximate 95% error limits were defined as bias ± 1.96 times the sample standard deviation of the vial-level errors; these limits describe error spread, not uncertainty in the mean bias.

Errors for iodine, gadolinium, tantalum, and bismuth were reported in mg/mL of the respective element; simulated-blood errors were reported in mg Fe/mL. Simulated-blood reference attenuation and tumor-model concentrations were reported as mean ± within-ROI spatial standard deviation. Tumor-model concentration errors were calculated relative to the prepared iodine or gadolinium concentration.

## Results

### Iodine and gadolinium quantification

Iodine and gadolinium were separated in both single-material and mixed vials (Figure 1). Two-input decomposition had the lowest iodine RMSE, 0.40 mg/mL, and the smallest absolute iodine bias, +0.01 mg/mL. Iodine RMSE was 0.73 mg/mL with three-material decomposition and 1.12 mg/mL with five-material decomposition.

Five-material decomposition had the lowest gadolinium RMSE, 0.46 mg/mL, compared with 0.55 mg/mL for three-material decomposition and 0.57 mg/mL for two-input decomposition. Three-material decomposition had the smallest absolute gadolinium bias, ™0.05 mg/mL, compared with ™0.08 mg/mL for five-material decomposition and +0.29 mg/mL for two-input decomposition (Figure 3; Table 1).

**Table 1.** Material-estimation errors in the common test vials.

Each cell reports signed bias / RMSE.
| Material output | Three-material decomposition | Five-material decomposition | Two-input decomposition |
| --- | --- | --- | --- |
| Iodine [mg I/mL] | −0.049 / 0.731 | +0.119 / 1.120 | +0.014 / 0.397 |
| Gadolinium [mg Gd/mL] | −0.054 / 0.554 | −0.082 / 0.459 | +0.291 / 0.572 |
| Simulated blood [mg Fe/mL] | +0.224 / 1.950 | −0.015 / 1.665 | — |
| Tantalum [mg Ta/mL] | — | +0.174 / 0.770 | — |
| Bismuth [mg Bi/mL] | — | −0.100 / 0.584 | — |

Bias was calculated as estimated minus reference concentration. The common test vials contained neither tantalum nor bismuth; therefore, the tantalum and bismuth rows quantify false-positive signal. A dash indicates that the material was not an output of that decomposition method.

**Figure 3.**
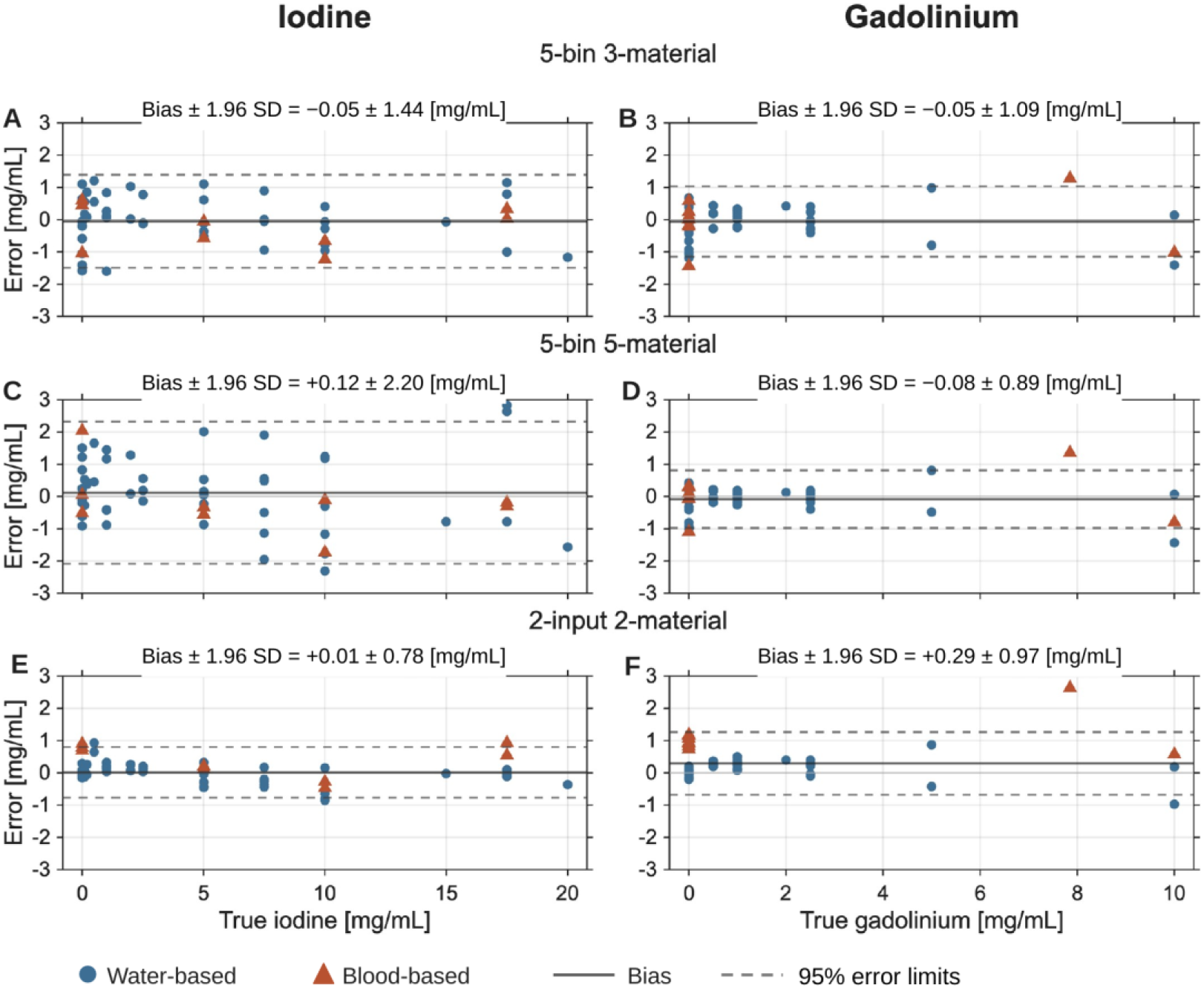
Iodine and gadolinium concentration errors for three material-decomposition methods. Panels A and B show three-material decomposition; C and D, five-material decomposition; and E and F, two-input decomposition. The first two methods use five energy-bin images, whereas the two-input method uses scanner-generated iodine and water images. The left and right columns show iodine and gadolinium, respectively. Each point represents the error for one vial, calculated as the mean estimated concentration minus the reference concentration. Blue circles indicate water-based preparations, and red triangles indicate preparations containing simulated blood. Solid lines show bias. Dashed lines show approximate 95% error limits, calculated as bias ± 1.96 times the sample standard deviation of vial-level errors. SD = standard deviation.

### Additional Material Separation

The simulated-blood reference measured 53.50 ± 10.20 HU at 70 keV. Five-material decomposition had a lower simulated-blood RMSE than three-material decomposition, 1.67 versus 1.95 mg Fe/mL, and a smaller absolute bias, ™0.02 versus +0.22 mg Fe/mL, respectively (Table 1).

Five-material decomposition also separated tantalum and bismuth. The tantalum test vial measured 9.69 mg Ta/mL, compared with the reference concentration of 10.00 mg Ta/mL. The bismuth test vial measured 10.82 mg Bi/mL, compared with the reference concentration of 10.10 mg Bi/mL. In test vials containing neither material, RMSE was 0.77 mg Ta/mL and 0.58 mg Bi/mL, with biases of +0.17 mg Ta/mL and ™0.10 mg Bi/mL, respectively.

### Tumor Vascular Models

Material maps separated the iodine-containing beads from gadolinium contrast within the tumor vascular models. The iodine-only and gadolinium-only controls showed signal predominantly in their corresponding material maps. In both bead-containing models, the iodine maps depicted the bead distribution, whereas the gadolinium maps depicted the contrast-filled vascular channels. Combined overlays showed the spatial relationship between the two materials within each model (Figure 2). Virtual monoenergetic images and individual material maps are shown in Figure 4.

**Figure 4.**
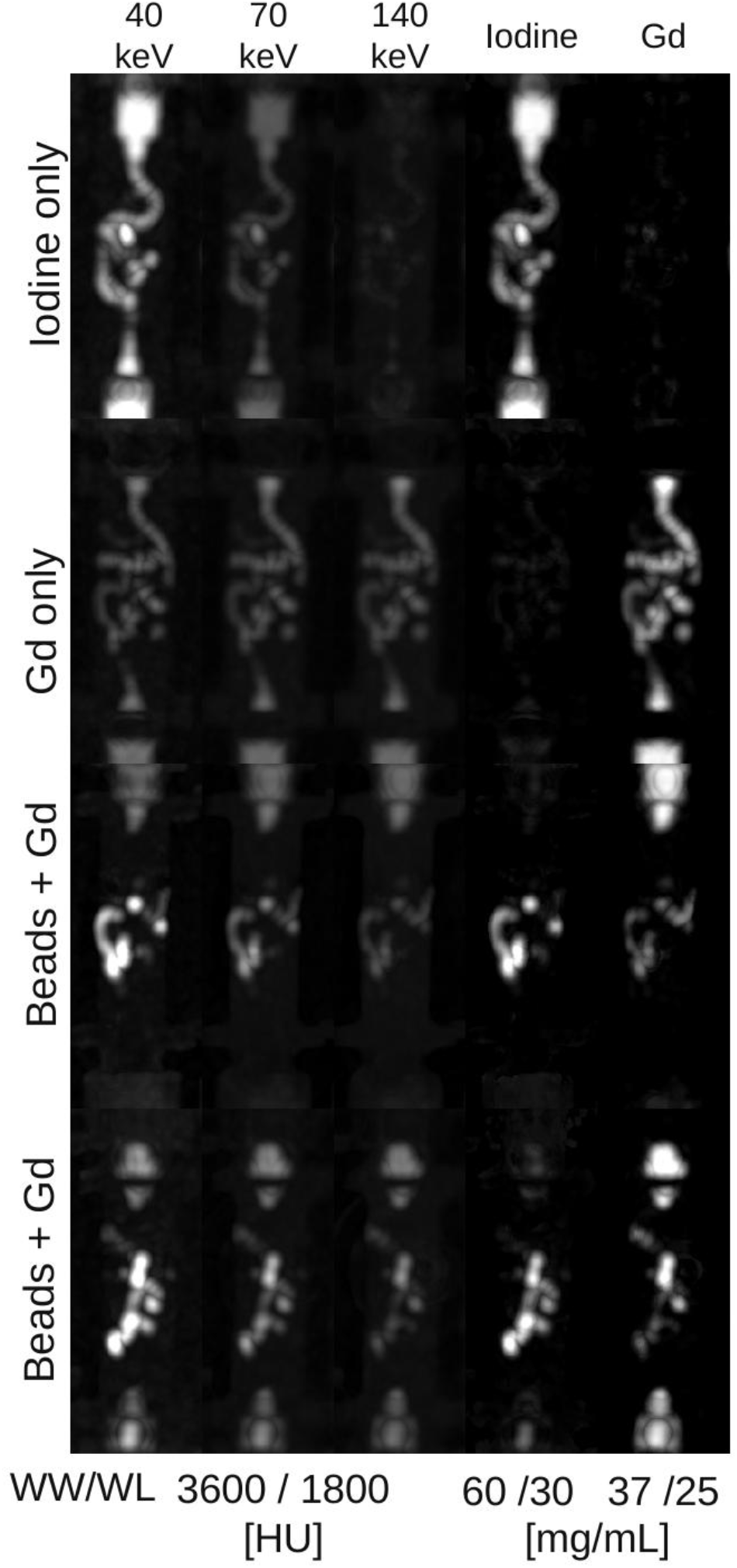
Virtual monoenergetic images and material maps of the tumor models. Columns show 40-, 70-, and 140-keV images, followed by iodine and gadolinium maps. Rows show the iodine-only control, gadolinium-only control, and two models containing radiopaque embolic beads and gadolinium. Images are z-axis maximum-intensity projections. Material maps were generated using two-input decomposition. WW/WL is 3600/1800 HU for the virtual monoenergetic images, 60/30 mg/mL for the iodine maps, and 37/25 mg/mL for the gadolinium maps. Gd = gadolinium; HU = Hounsfield units; WW/WL = window width/window level.

The iodine-only control measured 49.86 ± 8.52 mg I/mL, corresponding to an error of ™0.14 mg I/mL relative to the prepared concentration of 50.00 mg I/mL. The gadolinium-only control measured 37.58 ± 4.49 mg Gd/mL, corresponding to an error of +2.58 mg Gd/mL. In the two bead-containing models, gadolinium measured 32.56 ±8.02 and 37.02 ± 7.93 mg Gd/mL, corresponding to errors of ™2.44 and +2.02 mg Gd/mL, respectively, relative to the prepared concentration of 35.00 mg Gd/mL.

Cross-talk measured 2.71 ± 1.68 mg Gd/mL in the 50.00 mg I/mL iodine-only control and 2.47 ± 1.75 mg I/mL in the 35.00 mg Gd/mL gadolinium-only control.

## Discussion

Using deep-silicon photon-counting CT, we separated radiopaque embolic beads from gadolinium-filled vascular channels in 3D-printed tumor models and measured vascular gadolinium concentrations. Two-input decomposition of scanner-generated images provided quantitative iodine/gadolinium separation. Research decomposition of five reconstructed energy-bin images represented simulated blood separately and distinguished tantalum and bismuth. Together, these findings support the potential for dual-contrast post-embolization imaging and separation of additional materials.

Treatment assessment after embolization raises two spatial questions: where embolic material was deposited and whether enhancement persists within or around the treated region. In patients with hepatocellular carcinoma treated using radiopaque beads, bead density and residual tumor perfusion on cone-beam CT were associated with treatment response (Ruff et al., 2021). Enhancement also remains central to imaging assessment of viable treated tumor (Aslam et al., 2024). Separate material maps from one acquisition could display deposition and vascular contrast without registration to a second, unenhanced scan. This could be useful when motion or changes between examinations complicate subtraction. Our models demonstrate material separation, not the ability to determine tumor viability or measure residual blood flow.

Previous photon-counting CT studies used multiple contrast agents to distinguish enhancement phases in the heart and liver (Symons et al., 2017; Si-Mohamed et al., 2019) and contrast compartments in endoleak phantoms (Dangelmaier et al., 2018). Earlier deep-silicon work separated iodine and gadolinium in coronary phantoms (Holmes et al., 2024); a four-threshold research mode on a commercial photon-counting scanner has also quantified iodine and gadolinium in human-sized phantoms (Treb et al., 2025). The present application gives the materials different roles: iodine marks bead deposition, whereas gadolinium depicts vascular contrast. Their distinct spectral responses make this separation possible, but the present study does not compare scanner platforms.

The practical advantage of two-input decomposition is that it uses scanner-generated images with established vendor calibration and proprietary corrections. The scanner generates its iodine and water maps from all eight energy bins using projection-domain material decomposition. This estimates material composition from measured projection data before image reconstruction, accounting for the x-ray spectrum, detector response, and material-dependent beam hardening. Image-domain decomposition instead estimates material concentrations from reconstructed images and depends on the corrections applied during reconstruction (McCollough et al., 2020).

Direct comparisons have demonstrated better performance with projection-domain decomposition (Maaß et al., 2009). We subsequently calibrated the scanner-generated images to estimate iodine and gadolinium, using iodine and water images for the vial comparison and 70-keV and iodine images for the tumor models. Our research energy-bin decomposition used a linear image-domain model to demonstrate multimaterial feasibility. The accuracy comparison therefore reflects different processing implementations and material outputs.

Energy-bin images offer a different capability: more than two material responses can be represented separately. In our three-material model, the third output represented the attenuation of simulated blood. The five-material model additionally distinguished tantalum and bismuth, although each was evaluated in only one separate test vial. Prior photon-counting studies have demonstrated iodine, gadolinium, and bismuth imaging (Symons et al., 2017) and K-edge separation of other high-atomic-number materials in device and nanoprobe experiments (Brian-Choux et al., 2025; Gunaseelan et al., 2024). A separately estimated background could eventually support virtual noncontrast imaging that preserves unenhanced tissue, but this application was not tested here.

The simulated-blood mixtures tested whether contrast estimates remained accurate in an additional attenuating background. Water-only solutions cannot reveal this interference. In a two-output inverse, background signal may be assigned to iodine or gadolinium; a separately calibrated third output can represent some of that signal instead. Previous spectral micro-CT work separated iodine and gadolinium nanoparticle contrast agents in tumor-bearing mice (Badea et al., 2019). Our ferrous-sulfate preparation was an attenuation surrogate, not blood or perfused tissue, so the observed benefit of representing it separately needs validation in more realistic backgrounds.

The tumor models extended the vial measurements to an embolization-relevant geometry but remained static phantoms. Their concentrations represented local catheter-directed delivery rather than diagnostic intravenous enhancement. Mean gadolinium estimates in the two bead-containing models were near the prepared concentration, but the within-region spatial variation and cross-talk in single-material controls show that the maps were not perfectly specific. High local concentrations can cause material-dependent beam hardening (Pourmorteza et al., 2025); we did not isolate its contribution to the observed errors.

This phantom study demonstrates quantitative dual-contrast imaging and separation of radiopaque beads from vascular gadolinium under controlled conditions. These findings support future extension of calibrated multimaterial workflows beyond currently available material outputs. Perfused and in vivo studies are needed to determine whether these maps can assess residual enhancement after embolization under patient attenuation and motion.

## Data Availability

All data produced in the present study are available upon reasonable request to the authors.

## Declaration of generative AI and AI-assisted technologies in the manuscript preparation process

During the preparation of this work, the authors used OpenAI Codex to assist with manuscript organization and language revision, literature and citation verification, development of author-directed analysis code, and figure preparation. The authors defined the scientific questions and methods, supervised and verified the analyses, references, and figures, reviewed and edited the output as needed, and take full responsibility for the content of the published article.

